# Exposure to Heavy Metals, Obesity, and Stroke Mortality in the United States

**DOI:** 10.1101/2023.09.18.23295722

**Authors:** Ziqin Cao, Kelly M. Bakulski, Henry L. Paulson, Xin Wang

## Abstract

We investigated the associations between blood concentrations of lead and cadmium with stroke mortality, and potential effect modification by obesity. Our study analyzed data from 23,437 individuals aged 40 and above, using the National Health and Nutrition Examination Survey (NHANES 1999-2016) linked to the National Death Index. During a median follow-up period of 8.3 years, 336 stroke-related deaths were reported. After adjusting for potential confounders, we found that higher baseline concentrations of lead and cadmium were significantly associated with increased stroke mortality. Specifically, the hazard ratios (HRs) per doubling of metal concentrations were 1.16 (95% CI: 1.11, 1.20) for lead and 1.31 (95% CI: 1.26, 1.36) for cadmium. Stratified analysis showed that stronger associations were observed among participants who were normal weight or overweight, relative to those who were obese. In conclusion, our study demonstrates that elevated blood concentrations of lead and cadmium are significantly associated with an increased risk of stroke mortality, especially among individuals who are normal weight or overweight.

## 1. Introduction

Stroke is a major public health problem worldwide, it is a leading cause of death and disability globally, with an estimated 15 million new cases each year [1]. Despite significant advances in stroke management and prevention, the incidence and mortality rates of stroke remain high, underscoring the need for more effective prevention strategies. Identifying and targeting modifiable risk factors is crucial for characterizing high risk populations and identifying targets and strategies for prevention. Previous studies have identified lifestyle factors such as smoking, unhealthy diets, physical inactivity, and comorbidities like hypertension and diabetes as major risk factors for stroke [2-5]. While traditional risk factors have been well-established, there is increasing recognition of the role of environmental exposures in stroke risk.

Heavy metals, such as lead and cadmium, are ubiquitous environmental pollutants that have been linked to various adverse health outcomes, including an increased risk of cardiovascular disease and stroke. Biological evidence suggests that exposure to lead and cadmium induces oxidative stress, inflammation, and endothelial dysfunction, ultimately leading to atherosclerosis, increased blood pressure, and coagulation abnormalities, thereby elevating the risk of stroke [6-10]. At the same time, it is worth noting that persons with obesity have higher levels of oxidative stress, inflammation, and endothelial dysfunction [11-13]. Given the potential overlap in the biological mechanisms underlying obesity and the toxicity of heavy metals, it is possible that obesity may exacerbate the toxic effects of heavy metals on stroke risk. This highlights the need for further investigation into the potential interactions between these two types of risk factors, and the importance of identifying high-risk populations for targeted interventions.

The aim of this study is to investigate the association between exposure to heavy metals at baseline and stroke mortality during follow-up, and potential effect modification by obesity. We hypothesize that obesity may modify the relationship between heavy metals and stroke mortality, with obese individuals being more susceptible to the toxic effects of heavy metals. By addressing this research gap, we hope to provide valuable insights into the potential role of environmental exposures in stroke and to identify high-risk populations that may benefit from targeted prevention strategies.

## 2. Methods

### 2.1 Study Population

The study analyzed United States National Health and Nutrition Examination Survey (NHANES) data from nine 2-year cycles spanning 1999 to 2016, which used a stratified, multistage probability cluster design, with oversampling of selected subpopulations, to obtain a representative sample of the civilian, noninstitutionalized US population [14]. All data and materials have been made publicly available on the National Center for Health Statistics website (https://www.cdc.gov/nchs/nhanes/index.htm). The protocols for NHANES were approved by the National Center for Health Statistics of the Centers for Disease Control and Prevention Institutional Review Board, and informed consents were obtained from all participants. We defined our study baseline as the NHANES exposure measure, and we used linkage to the National Death Index for follow-up outcome measures.

The study population included adults aged 40 years or older who had blood lead and cadmium measured (n=32,366). We restricted to adults aged 40 years or older because participants younger than 40 have very low cardiovascular and cerebrovascular disease risks. Including these low-risk participants may skew the model and result in inaccurate predictions [15-16]. Of these adults, we further excluded participants without linkable mortality data in the National Death Index (n=54) or missing data on blood lead/cadmium (n=6536), cholesterol (n=3478), body mass index (n=2405), smoking (n=51), and blood pressure (n=3486). The final analytic sample included 23,437 participants (**Figure 1**).

**Figure 1.**
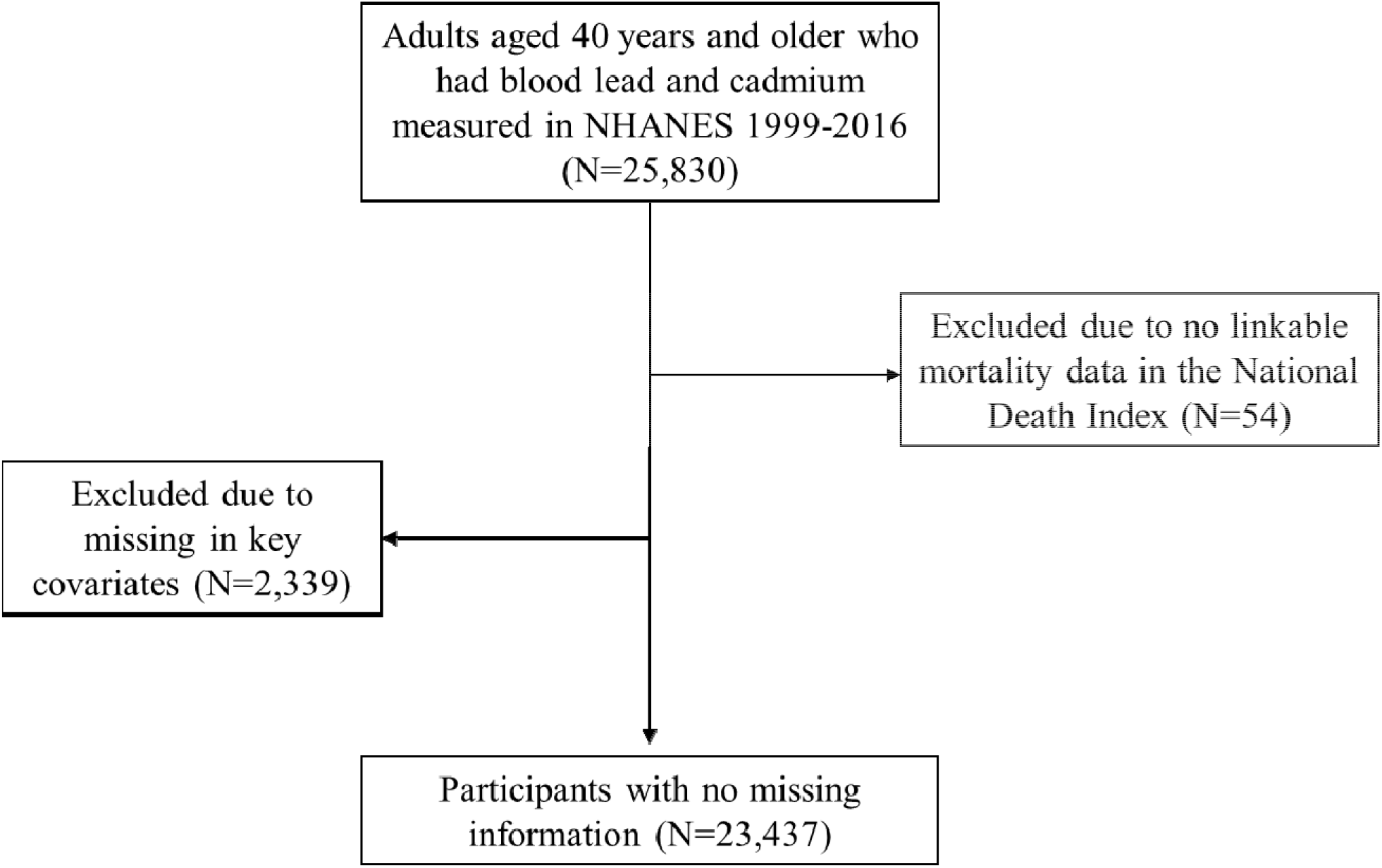
Flow chart depicting included and excluded participants from the National Health and Nutrition Examination Survey (NHANES) 1999-2016.

### 2.2 Stroke mortality

Vital status and cause of death were determined using the publicly available NHANES-linked mortality data up to December 31, 2019. Probabilistic matching algorithms were used to link NHANES records with death certificates from the National Death Index. Further information regarding the matching criteria and calibration can be found in the National Center for Health Statistics documentation [17]. Death from cerebrovascular causes was determined based on the following International Classification of Disease-10 codes (ICD-10): I60-I69.

### 2.3 Blood lead and cadmium measurements

Lead and cadmium were measured in whole blood simultaneously during the 1999-2002 survey years using atomic absorption spectrophotometry [18-21]. For the survey years of 2003-2016, inductively coupled plasma mass spectrometry (ICP-MS) was used to determine whole blood lead and cadmium concentrations, which is a multi-element analytical technique based on quadrupole ICP-MS technology [22]. The lower detection limits (LOD) for lead were 0.30 μg/dL for NHANES 1999-2004 and 0.25 μg/dL for 2005-2016, while for cadmium, LOD was 0.30 μg/dL for NHANES 1999-2002, 0.20 μg/dL for 2003-2010, and 0.16 μg/dL for 2011-2016. Results below the LOD were replaced with a value equal to LOD divided by the square root of two [23].

### 2.4 Covariate measures

Data on participants’ age, sex, race/ethnicity (including Hispanic and Mexican American, non-Hispanic white, non-Hispanic black, and others), as well as smoking status, were collected through self-filled questionnaires. Blood pressure measurements were taken three or sometimes four times for each participant while seated, following a standard procedure based on the American Heart Association guidelines. Average systolic blood pressure was determined by taking the mean of up to three readings, excluding the initial one. Hypertension was then defined as systolic blood pressure ≥140 mmHg, or diastolic blood pressure ≥90 mmHg, or usage of antihypertensive medications. Total cholesterol levels in the serum (mg/dL) were assessed using enzymatic assays. BMI was calculated as the weight in kilograms divided by the square of the height in meters. The criteria for diabetes included either a self-reported diagnosis, usage of antidiabetic medications, or a hemoglobin A1c level of 6.5% or higher.

### 2.5 Statistical analysis

Mean and standard deviation (SD) were computed for continuous variables, and percentage (%) was calculated for categorical variables. Participant characteristics and blood metal concentrations were compared by stroke mortality status.

Survey-weighted Cox Proportional Hazards models were used to evaluate the associations between blood metal concentrations and stroke mortality. Participants contributed survival time from the NHANES examination to the date of death for those who died from stroke and participants who died from other causes were right-censored at the date of death, and those who were alive were right-censored at the last follow-up date (December 31, 2019). Given the right-skewed distributions of metal concentrations, logarithmic transformations with base two were applied to all metal concentrations. Effect estimates were interpreted as hazard ratios (HR) of stroke death per doubling of each blood metal concentration. The associations between metals and stroke mortality were further stratified by obesity status (BMI>=30, 25-30, <25). All models were adjusted for age, sex, race/ethnicity (Non-Hispanic White, Non-Hispanic Black, Mexican American, Other Hispanic, and other racial/ethnic groups), smoking (never smoker, former smoker, and current smoker), total cholesterol, diabetes (yes vs no), and hypertension (yes vs no). The sampling weights and design variables were used for all regression models to generalize the findings to the U.S. general population. All analyses were conducted using R, version 4.2.1 (www.R-project.org).

## 3. Results

**Table 1** presents the baseline characteristics of the study cohort. The total population consisted of 23,437 individuals, among whom 336 individuals died from stroke and 23,101 individuals were non-stroke death or alive. The mean age of the study population was 59.9 years (SD=12.7), and the median follow-up time was 10.1 years.

**Table 1.** Baseline Characteristics of the Study Cohort.

|  | **Total population**<br>**(N=23,437)** | **Non-stroke death or**<br>**alive (N=23,101)** | **Stroke death**<br>**(N=336)** |
| --- | --- | --- | --- |
| Blood lead, mean (SD),<br>$\mu\text{g/dL}$ | 1.67 (1.41) | 1.66 (1.40) | 2.10 (1.65) |
| Blood cadmium, mean (SD),<br>$\mu\text{g/dL}$ | 0.40 (0.41) | 0.40 (0.40) | 0.49 (0.38) |
| BMI, No. (%) |  |  |  |
| $\geq 30 \text{ kg/m}^2$ | 8,726 (37.2) | 8,620 (37.3) | 106 (31.6) |
| $\geq 25$ and $< 30 \text{ kg/m}^2$ | 8,557 (36.5) | 8,423 (36.5) | 134 (39.9) |
| $< 25 \text{ kg/m}^2$ | 6,154 (26.3) | 6,058 (26.2) | 96 (28.6) |
| Age, mean (SD), year | 59.9 (12.7) | 59.7 (12.6) | 72.2 (10.1) |
| Female, No. (%) | 11,756 (50.2) | 11,598 (50.2) | 158 (47.0) |
| Racial/Ethnic groups, No.<br>(%) |  |  |  |
| Hispanic<sup>†</sup> | 4,003 (17.1) | 3,953 (17.1) | 50 (14.9) |
| Non-Hispanic White | 11,631 (49.6) | 11,435 (49.5) | 196 (58.3) |
| Non-Hispanic Black | 4,556 (19.4) | 4,500 (19.5) | 56 (16.7) |
| Other racial/ethnic groups | 1,463 (6.2) | 1,449 (6.3) | 14 (4.2) |
| Smoking status, No. (%) |  |  |  |
| Current smoker | 4,424 (18.9) | 4,370 (18.9) | 54 (16.1) |
| Former smoker | 7,394 (31.6) | 7,265 (31.5) | 129 (38.4) |
| Never smoker | 11,619 (49.6) | 11,466 (49.6) | 153 (45.5) |
| Total Cholesterol, mean (SD), mg/dL | 202.7 (42.5) | 202.7 (42.4) | 200.4 (43.1) |
| Diabetes, No. (%) | 4,034 (17.2) | 3950 (17.1) | 84 (25.0) |
| Hypertension, No. (%) | 10,881 (46.4) | 10657 (46.1) | 224 (66.7) |

\* Non-stroke death included both alive and death of other causes.
<sup>†</sup> Combined from Mexican American and other Hispanic in NHANES.

In terms of blood metal concentrations, the mean blood lead concentration was slightly higher in the stroke death group (2.10 μg/dL, SD=1.65) compared to the non-stroke death or alive group (1.66 μg/dL, SD=1.40). The mean blood cadmium concentration was also slightly higher in the stroke death group (0.49 μg/dL, SD=0.38) compared to the non-stroke death or alive group (0.40 μg/dL, SD=0.40).

Regarding other characteristics, there were slightly more individuals with a BMI ≥30 in the non-stroke death or alive group (37.3%) compared to the stroke death group (31.5%). The stroke death group had a higher proportion of individuals with hypertension (66.7%) and diabetes (25.0%) compared to the non-stroke death or alive group (46.1% and 17.1%, respectively).

**Table 2** shows the median (IQR) metal concentrations in μg/dL by participant characteristics. A higher median lead concentration was observed among Non-Hispanic Black participants, current smokers, and those with a BMI below 25. Participants from other racial/ethnic groups and those who are current smokers showed higher median cadmium concentrations.

**Table 2.** Metal Concentrations by Participant Characteristics.

| Characteristic | Metal concentrations, Median (IQR), µg/dL |  |
| --- | --- | --- |
|  | Lead | Cadmium |
| BMI | 1.50 (1.22) | 0.38 (0.36) |
| ≥30 | 1.76 (1.49) | 0.40 (0.36) |
| ≥25 and <30 | 1.84 (1.60) | 0.50 (0.58) |
| <25 | 1.50 (1.22) | 0.38 (0.36) |
| Age, year |  |  |
| 40-59 | 1.49 (1.24) | 0.40 (0.48) |
| ≥ 60 | 1.89 (1.54) | 0.42 (0.34) |
| Sex |  |  |
| Female | 1.40 (1.40) | 0.42 (0.38) |
| Male | 2.00 (1.65) | 0.39 (0.43) |
| Racial/Ethnic groups |  |  |
| Hispanic* | 1.64 (1.50) | 0.40 (0.36) |
| Non-Hispanic White | 1.70 (1.38) | 0.40 (0.41) |
| Non-Hispanic Black | 1.80 (1.68) | 0.40 (0.48) |
| Other racial/ethnic groups | 1.56 (1.27) | 0.53 (0.57) |
| Smoking status |  |  |
| Current smoker | 2.15 (1.73) | 1.05 (0.88) |
| Former smoker | 1.80 (1.48) | 0.40 (0.32) |
| Never smoker | 1.42 (1.20) | 0.31 (0.27) |
| Diabetes |  |  |
| Yes | 1.43 (1.31) | 0.39 (0.37) |
| No | 1.70 (1.46) | 0.40 (0.44) |
| Hypertension |  |  |
| Yes | 1.70 (1.48) | 0.40 (0.40) |
| No | 1.63 (1.40) | 0.40 (0.42) |

\* Combined from Mexican American and other Hispanic in NHANES.

**Table 3** shows the HR with 95% confidence intervals (CI) for stroke death in relation to each metal concentration. In the total population, the HR for stroke death per doubling of lead concentration is 1.16 (95% CI: 1.11, 1.20), and for cadmium is 1.31 (95% CI: 1.26, 1.36), after adjusting for age, sex, race/ethnicity, smoking, total cholesterol, diabetes, and hypertension.

**Table 3.**
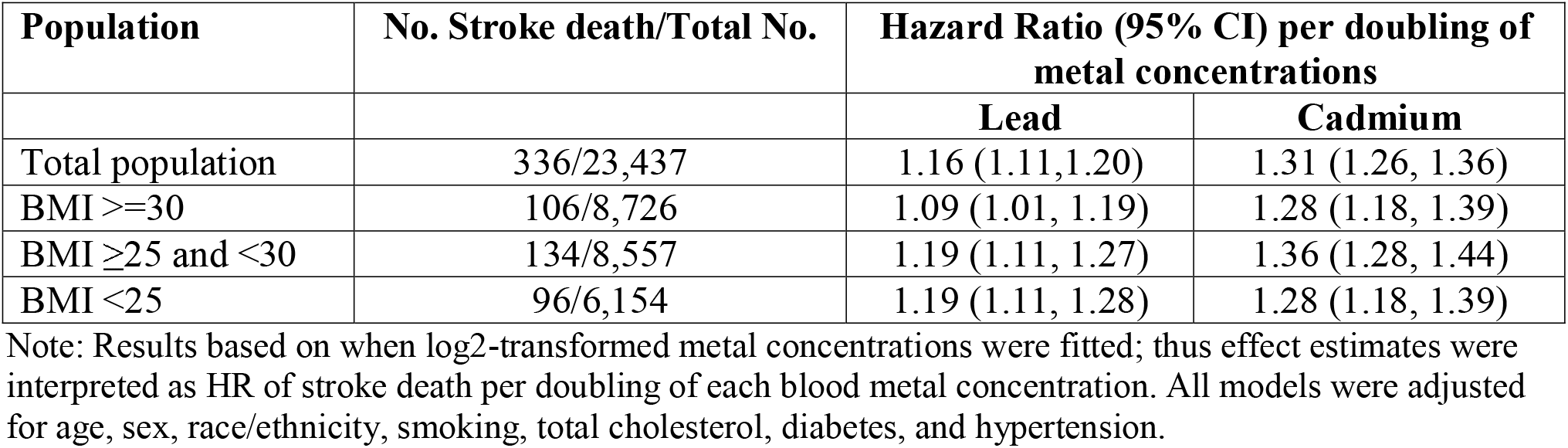
Hazard ratios (HR) (95% confidence intervals, 95% CI) for incident metabolic syndrome in relation to blood metal concentrations.

| Population | No. Stroke death/Total No. | Hazard Ratio (95% CI) per doubling of metal concentrations |  |
| --- | --- | --- | --- |
|  |  | Lead | Cadmium |
| Total population | 336/23,437 | 1.16 (1.11, 1.20) | 1.31 (1.26, 1.36) |
| BMI $\geq 30$ | 106/8,726 | 1.09 (1.01, 1.19) | 1.28 (1.18, 1.39) |
| BMI $\geq 25$ and $< 30$ | 134/8,557 | 1.19 (1.11, 1.27) | 1.36 (1.28, 1.44) |
| BMI $< 25$ | 96/6,154 | 1.19 (1.11, 1.28) | 1.28 (1.18, 1.39) |

Note: Results based on when log2-transformed metal concentrations were fitted; thus effect estimates were interpreted as HR of stroke death per doubling of each blood metal concentration. All models were adjusted for age, sex, race/ethnicity, smoking, total cholesterol, diabetes, and hypertension.

In stratified analyses by BMI categories (obese, BMI ≥ 30; overweight, 25 ≤ BMI < 30; and normal, BMI < 25), the association between metal concentrations and stroke death remained significant in each group. The HRs for both lead and cadmium concentrations are consistently higher in the overweight and normal BMI categories than in the obese BMI category.

## 4. Discussion

In this study, we investigated the associations of blood lead and cadmium with stroke mortality in a large, diverse, longitudinal sample of US adults using the NHANES. Higher concentrations of blood lead and cadmium were associated with elevated stroke mortality. These associations persisted after adjusting for conventional cardiovascular risk factors. We further stratified associations by BMI categories, and stronger associations were observed among participants who were overweight or had a normal BMI.

Previous studies have also reported positive associations between metal exposure and stroke mortality. In a prospective cohort study of elderly men, Weisskopf et al. reported a strong positive association between bone lead concentration and the risk of ischemic stroke [24]. Another paper by Tellez-Plaza, Maria et al. also found strongly suggestive evidence that cadmium, at substantially low levels of exposure, remains an important determinant of all-cause and cardiovascular disease mortality in a representative sample of U.S. adults [25]. The findings suggest that efforts to reduce exposure to lead and cadmium may have important public health implications for reducing the risk of stroke mortality.

In stratified analysis, stronger associations of lead and cadmium with stroke mortality were observed, in contrast to our hypothesis. However, to the best of our knowledge, there is no biological evidence supporting that obesity itself would induce cerebrovascular benefits against heavy metals’ toxicity. The observation of a stronger association among non-obese individuals may also suggest the presence of an “obesity paradox” in the association between metals and stroke mortality, where the negative health effects of obesity may be partially offset by protective factors [26-28]. Additionally, obese populations would have a higher profile of other confounding factors, such as hypertension, diabetes, and dyslipidemia, which may mask the direct effects of lead and cadmium exposure on stroke mortality. Nevertheless, further research is required to elucidate the complex interplay between obesity, toxic metal exposure, and stroke risk.

The underlying biological mechanisms that link metal exposure to stroke mortality are not fully understood, but several possible mechanisms have been proposed. For example, lead exposure may lead to oxidative stress and inflammation, which can damage the vascular endothelium and contribute to atherosclerosis. Similarly, cadmium exposure may promote oxidative stress, endothelial dysfunction, and inflammation, leading to cardiovascular damage [29-33]. These pathways may explain the observed association between metal exposure and stroke mortality. In the case of the stronger association observed among overweight individuals, it is possible that obesity may exacerbate these mechanisms by promoting inflammation and endothelial dysfunction, which could increase the negative health effects of metal exposure.

The present study has several strengths, including the use of a representative sample of the US population, the adjustment for several potential confounding variables, and the ability to examine the associations stratified by obesity. However, there are also several limitations to consider. First, the study only considered stroke mortality and not incident cases of stroke, which may limit the generalizability of the findings to overall stroke risk. Second, the study only examined lead and cadmium exposure and did not consider other metals that may also be relevant to stroke risk. Third, as with any observational study, there may be unmeasured confounding factors that could influence the observed associations. Nonetheless, the findings of this study provide important insights into the potential health effects of metal exposure and underscore the need for continued efforts to reduce exposure to harmful environmental toxcants.

## 5. Conclusion

In summary, our study found that metal lead and cadmium concentrations were associated with an increased risk of stroke mortality over eight years of follow-up, particularly among normal weight and overweight individuals. These findings highlight the importance of reducing exposure to lead and cadmium to reduce the risk of stroke mortality. Additional studies with prospective cohort designs and incidence data are needed to confirm these findings. Furthermore, more research is needed to determine whether interventions to reduce metal exposure can effectively reduce the risk of stroke mortality.

## Acknowledgement

The authors would also like to thank the NHANES participants and the staff members for their contribution to data collection and for making the data publicly available.

## Data availability

All data and materials have been made publicly available at the National Center for Health Statistics website (https://www.cdc.gov/nchs/nhanes/index.htm).

